# Glutathione as a molecular marker of functional impairment in patients with at-risk mental state: 7-Tesla ^1^H-MRS study

**DOI:** 10.1101/2020.11.17.20233635

**Authors:** Peter Jeon, Roberto Limongi, Sabrina D. Ford, Cassandra Branco, Michael Mackinley, Maya Gupta, Laura Powe, Jean Théberge, Lena Palaniyappan

## Abstract

A substantial number of individuals with clinical high-risk (CHR) mental state do not transition to psychosis. However, regardless of future diagnostic trajectories, many of these individuals develop poor social and occupational functional outcomes. The levels of glutathione, a crucial cortical antioxidant, may track variations in functional outcomes in early psychosis and prodromal states.

Thirteen clinical high-risk and 30 healthy control volunteers were recruited for a 7-Tesla magnetic resonance spectroscopy scan with voxel positioned within the dorsal anterior cingulate cortex (ACC). Clinical assessment scores were collected to determine if any association was observable with glutathione levels.

Bayesian Spearman test revealed a positive association between the Social and Occupational Functioning Assessment Scale (SOFAS) and the glutathione concentration in the clinical high-risk group but not in the healthy control group. After accounting for variations in SOFAS, CHR group had higher GSH levels than the healthy subjects.

This study is the first to use 7-Tesla magnetic resonance spectroscopy to test whether ACC glutathione levels related to social and occupational functioning in a clinically high-risk group and offers preliminary support for glutathione levels as a clinically actionable marker of prognosis in emerging adults presenting with risk features for various severe mental illnesses.

## Introduction

Emerging adults with attenuated or brief and limited psychotic symptoms are said to be in a clinical high-risk (CHR) state (or “at-risk or ultra high-risk” mental state) that later develops into multiple diagnostic outcomes including schizophrenia, mood disorders such as bipolar disorder or major depressive disorder [1,2]. A substantial number of individuals with CHR develop poor long-term functional (i.e., social and occupational) outcomes, irrespective of diagnostic transitions. Longitudinal studies indicate that a large proportion of individuals with CHR do not transition to psychosis (65-89% not psychotic over 2-10 years [3–5]), but have poor social and occupational outcomes (48% functionally impaired at 3-10 years [3,6]). While functional outcomes improve over time in CHR patients who have good functioning at the baseline, persistent deficits are seen in those who start with lower levels of functioning [4]. In other words, lower levels of functioning at the CHR state, before the onset of diagnosable psychiatric disorders such as schizophrenia and bipolar disorder, reliably predicts the trajectory of continued poor functioning over a long time period. The molecular bases of such pervasive functional deficits continue to be unknown [7], proving to be a major hurdle in developing meaningful treatments aimed at the CHR state.

Oxidative stress has emerged as a key mechanism underlying the pathophysiology of many psychiatric disorders including psychosis [8]. Destructive free radicals that damage brain tissue are by-products of oxidative metabolism but are effectively scavenged by antioxidants. Glutathione (GSH), the cardinal antioxidant in brain cells, shows 27-52% reduction [9–11] in established schizophrenia. Genetic [12,13] and cell biology studies [14–16] indicate that in a subset of patients, GSH production on demand is likely to be reduced [17]. We recently demonstrated the prognostic importance of low GSH in predicting early clinical response to antipsychotics in first episode schizophrenia. In this study, we observed that for every 10% baseline difference in anterior cingulate cortex (ACC) GSH among patients, 7 additional days of delay in response occurred after treatment initiation [18]. Lack of early response is a critical indicator of long-term poor outcomes in schizophrenia [19–21]. We and others have also related lower GSH to various determinants of functional outcomes including residual symptom burden [22], negative symptoms [23] and cognitive deficits [24], supporting the notion that the “hub of oxidative stress” indexed by GSH [25,26] is likely a critical determinant of functioning.

We recently synthesized in vivo magnetic resonance spectroscopy (MRS) studies in the ACC and demonstrated a significant GSH reduction in established cases of schizophrenia but an elevation in bipolar disorder [27], indicating that GSH levels may track the variations in functional outcomes that typify the prognostic course of psychiatric disorders. Such divergence between disorders may mean that in the ‘pluripotent’ CHR state that includes patients with varying levels of functioning as a single group, GSH levels may not differ from healthy controls, but will relate to variations in levels of functioning. In fact, in the only previous MRS study of cortical glutathione in clinical high-risk state [28], da Silva and colleagues reported no difference between healthy controls and CHR subjects in anterior cingulate glutathione [29]. Functional outcomes were not evaluated in this study; thus, the role of GSH as a transdiagnostic prognostic marker in CHR remains unknown.

In the current study, we use ultrahigh field 7T MRS for the first time to test if ACC GSH levels relate to social and occupational functioning in the CHR group. We expected GSH levels to be reduced among patients with poor functioning. Furthermore, we aimed at establishing the difference in GSH levels between CHR and healthy control subjects after accounting for variations in functioning. We evaluated these hypotheses using a Bayesian statistical approach.

## Results

### Demographic Data

Subject demographic and clinical data are summarized in Table 1. A small number within the clinical high-risk group were being administered antidepressant (N = 3) or benzodiazepine (N = 2) at the time of scan. Mean percent CRLB values for CHR and HC GSH were 10% ± 4% and 11% ± 4%, respectively. CHR patients had substantially high levels of functional impairment. Groups differed in CAST scores, being higher in the CHR group than in the HC group (mode = 4.19, posterior proportion [PP] = 1.0) but not in AUDIT-C scores (mode = -0.01, PP = 0.6). SOFAS scores were higher in the HC group than in the CHR group (mode = 16.7, PP = 1.0).

**Table 1.**
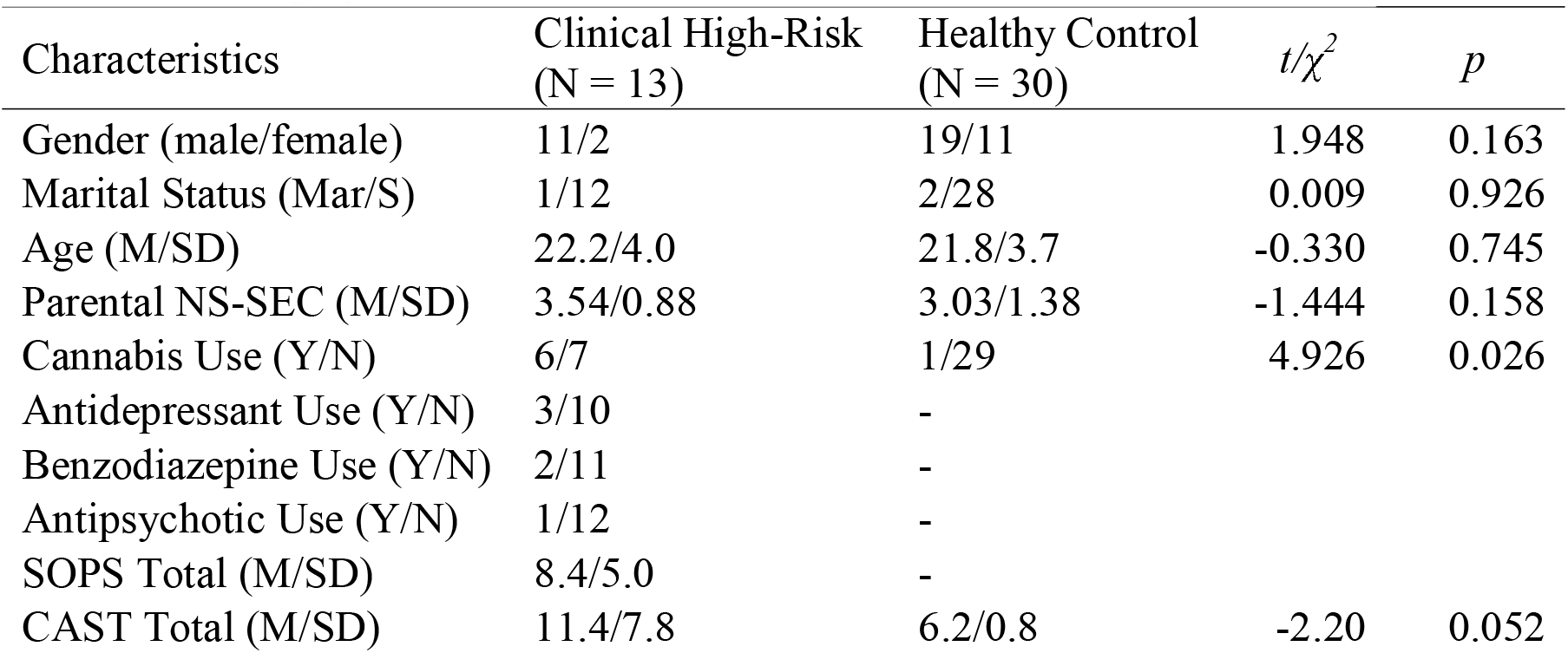
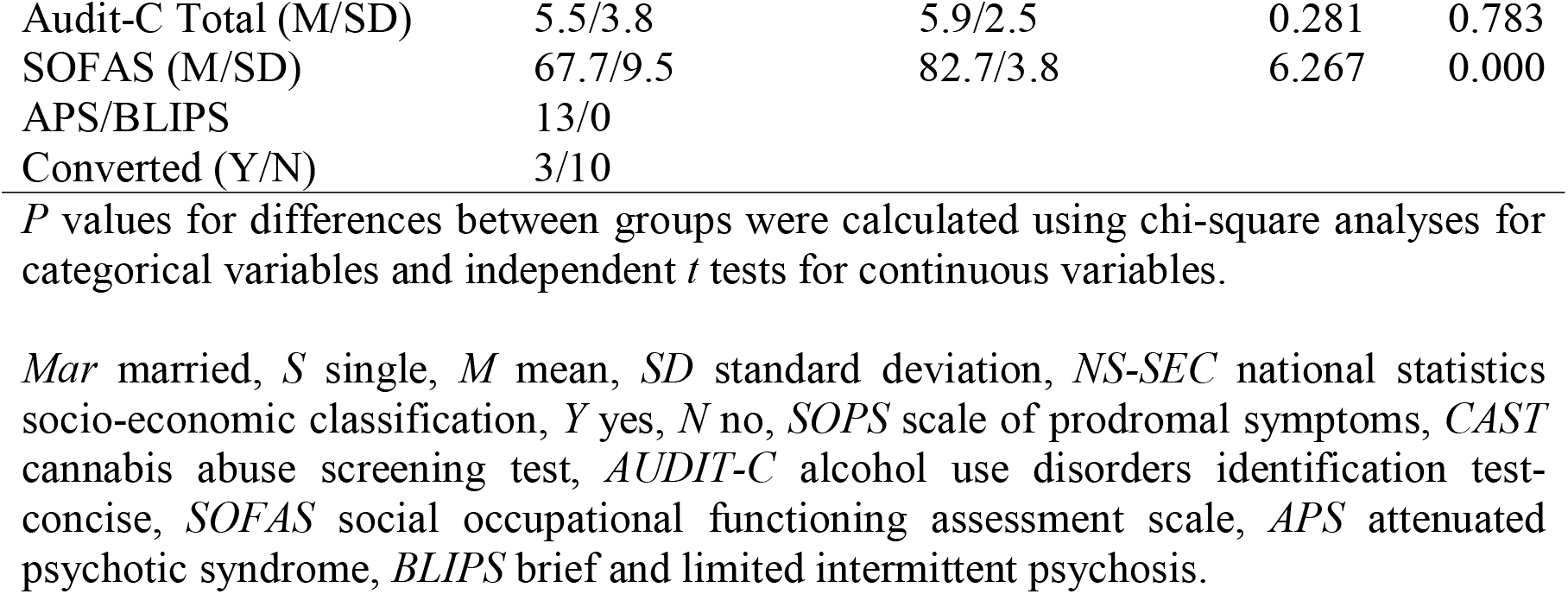
Demographics and clinical characteristics.

### GSH, CHR Status and Social and Occupational Functioning

The Spearman test revealed a positive association between the Social and Occupational Functioning Assessment Scale (SOFAS) scores and the [GSH] in the CHR group (mode ρ = 0.58, posterior proportion [PP] = 0.98, Bayesian Factor in favour of H_1_ over the null H_0_ [BF_10_] = 2.1) whereas, in the HC, the test speaks to “absence of effect” (mode ρ = 0.11, PP = 0.44, BF_10_ =0.23). In the CHR group, there was neither effect of SOPS (mode ρ = -0.17, PP = 0.78, BF_10_ = 0.22) nor effect of CAST (mode ρ = 0.32, PP = 0.87, BF_10_ = 0.46) on [GSH] (Table 2).

**Table 2.**
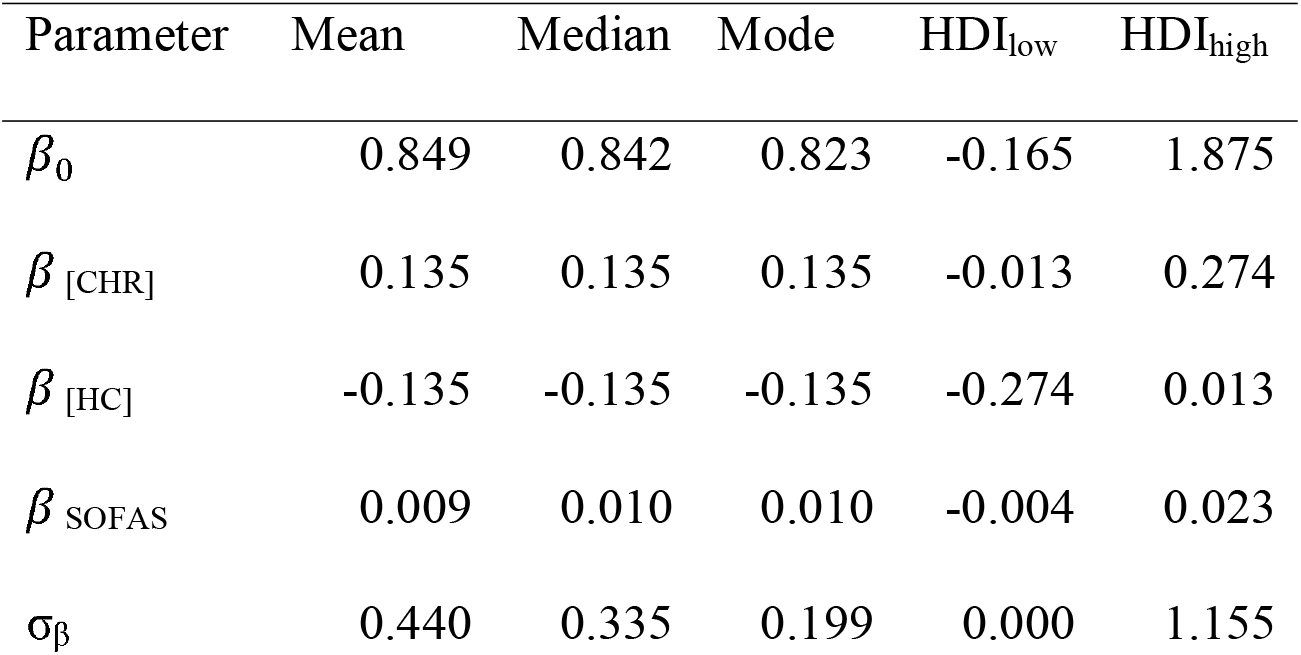
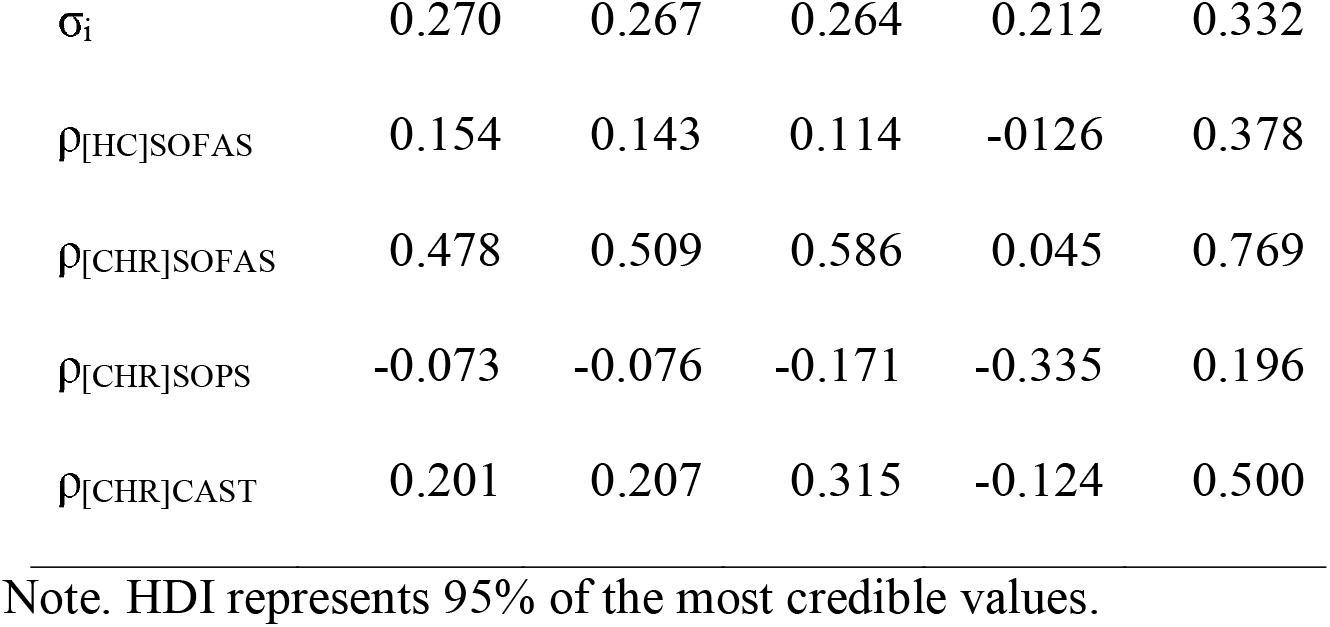
Parameter estimates (Posteriors) of the hierarchical Bayesian linear model and the Spearman’s correlation test.

| Parameter | Mean | Median | Mode | HDI<sub>low</sub> | HDI<sub>high</sub> |
| --- | --- | --- | --- | --- | --- |
| $\beta_0$ | 0.849 | 0.842 | 0.823 | -0.165 | 1.875 |
| $\beta_{[CHR]}$ | 0.135 | 0.135 | 0.135 | -0.013 | 0.274 |
| $\beta_{[HC]}$ | -0.135 | -0.135 | -0.135 | -0.274 | 0.013 |
| $\beta_{SOFAS}$ | 0.009 | 0.010 | 0.010 | -0.004 | 0.023 |
| $\sigma_\beta$ | 0.440 | 0.335 | 0.199 | 0.000 | 1.155 |
| $\sigma_i$ | 0.270 | 0.267 | 0.264 | 0.212 | 0.332 |
| $\rho_{[HC]SOFAS}$ | 0.154 | 0.143 | 0.114 | -0.126 | 0.378 |
| $\rho_{[CHR]SOFAS}$ | 0.478 | 0.509 | 0.586 | 0.045 | 0.769 |
| $\rho_{[CHR]SOPS}$ | -0.073 | -0.076 | -0.171 | -0.335 | 0.196 |
| $\rho_{[CHR]CAST}$ | 0.201 | 0.207 | 0.315 | -0.124 | 0.500 |

Note. HDI represents 95% of the most credible values.

*HDI* highest density interval, *β*_0_ intercept, *β* _[CHR]_ deflection parameter for the CHR group, *β* _[HC]_ deflection parameter for the HC group, *β*_SOFAS_ deflection parameter for the SOFAS covariate,σ_β_ standard deviation of the baseline parameter,σ_i_ standard deviation of the predicted value, ρ_[HC]SOFAS_ Spearman’s correlation between SOFAS score and GSH of control group, ρ_[CHR]SOFAS_ Spearman’s correlation between SOFAS score and GSH of CHR group, ρ_[CHR]SOPS_ Spearman’s correlation between SOPS score and GSH of CHR group, ρ_[CHR]CAST_ Spearman’s correlation between CAST score and GSH of CHR group.

After accounting for the SOFAS scores, the metabolite level in the HC group was smaller than in the CHR group (mode difference = -0.26, PP = 0.96; effect size -1.04, PP = 0.96). Summary statistics of the posterior distributions of the model’s parameter estimates are reported in Table 2. Figure 1 shows the posterior distributions of the estimated between-groups difference in GSH. For completeness, a frequentist analysis is presented in Supplementary Materials.

**Figure 1.**
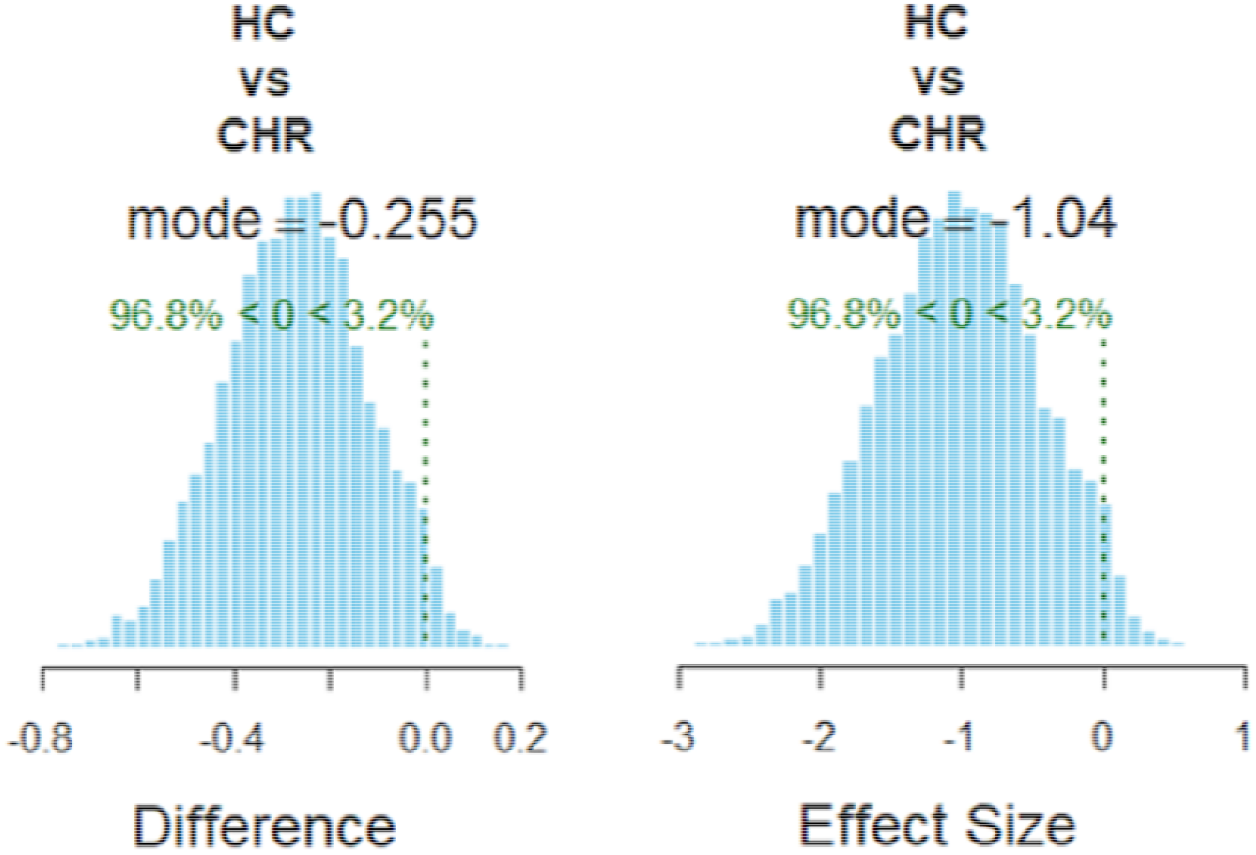
Bayesian analysis. Poster distributions of the estimated between-groups difference in glutathione.

## Discussion

Our data provides evidence in support of a relationship between GSH levels and social and occupational functioning in clinical high-risk state, and the presence of higher GSH in CHR subjects, when the variance related to functional impairment is accounted for. In this study, we observed no relationship between GSH levels in the ACC and SOFAS in healthy volunteers, especially as the functional variability was within a narrow range among the healthy subjects. Furthermore, we did not observe any correlations between GSH and prodromal positive symptom severity. Taken together, these results support our hypothesis that GSH is a key molecular substrate underlying the functional deficits seen in CHR state.

An exciting translational utility of identifying the GSH-deficit in low functioning patients is the therapeutic possibility of correcting it. A number of compounds with the potential to correct th effects of GSH deficit are in the pipeline [30–32]. Of these, N-acetylcysteine has been shown to improve cognition and negative symptoms in schizophrenia (6 RCTs) [33], and global functioning in mood disorders [34,35]. Given the persistent nature of functional deficits, reversing them will likely requires longer trials that are substantially difficult to complete. Antioxidants that increase GSH levels are more likely to benefit patients whose GSH levels are lower to begin with [36]. Our results support stratifying antioxidant trials on the basis of baseline functional impairment or GSH levels in the future.

Our study has a number of strengths. We used 7T-MRS sequence with improved specificity to detect GSH resonance with reduce macromolecular interference [18,37]. Among the MRS studies specifically optimized for GSH detection, 7T studies [22,38] report higher effect sizes for GSH reduction in schizophrenia compared to 3T [12,39]. We also recruited patients who were not treated with antipsychotics, and evaluated an age, gender and parental socioeconomic status matched control group. Nevertheless, our sample size was limited compared to the prior study addressing this question using a 3T-MRS sequence. Furthermore, we lacked the follow-up data necessary to identify transition to psychosis among the CHR groups. From the published meta-analytical data, we expect 1-3 converters in the next 2-5 years of observation [40].

## Materials and Methods

### Participants

We recruited 13 clinical high-risk (CHR) volunteers along with 30 healthy control (HC) volunteers, group-matched for age, gender, and parental socio-economic status. Patient volunteers were recruited from the referrals received by the PROSPECT (Prodromal Symptoms of Psychosis – Early Clinical Identification and Treatment) program at London Health Sciences Center, London, Ontario. Patients were help-seeking individuals referred to the clinic by community physicians, healthcare workers or friends/family. All referrals were reviewed by an intake coordinator via telephone using a validated instrument [PRIME Screen – Revised]. If found eligible for further assessment, the patients were evaluated within 2 weeks of referral using Structured Interview for Psychosis-risk Syndromes (SIPS) [41]. Patients with medical conditions, pervasive developmental disorders or intellectual disability underlying the reported symptoms, those who received treatment with antipsychotic medications to treat presenting symptoms (minimal effective dose for a period of at least 2 weeks), and those with psychotic symptoms secondary to active substance use (intoxication effects) were excluded. Based on SIPS, patients satisfying Attenuated Psychotic Syndrome (APS) or Brief and Limited Intermittent Psychosis (BLIPS) were both included in the CHR group. Healthy volunteers had no personal history of mental illness with no family history of psychotic disorder. All participants were screened to exclude significant head injury, major medical illness, or MRI contraindications and provided written, informed consent according to the guidelines of the Human Research Ethics Board for Health Sciences at Western University, London, Ontario.

### MRS Acquisition and Analysis

A Siemens MAGNETOM 7T head-only MRI scanner (Siemens, Erlangen, Germany) was used for all MRS acquisition along with a site-built head coil (8-channel transmit, 32-channel receive) at the Centre for Functional and Metabolic Mapping of Western University (London, Ontario). A two-dimensional sagittal anatomical image (37 slices, TR = 8000 ms, TE = 70 ms, flip-angle (α) = 120°, thickness = 3.5 mm, field of view = 240×191 mm) was used as reference to prescribe a 2.0 x 2.0 x 2.0 cm (8 cm^3^) ^1^H-MRS voxel on the bilateral dorsal ACC (Figure 2). Voxel position was prescribed by setting the posterior face of the voxel to coincide with the precentral gyrus and setting the position of the inferior face of the voxel to the most caudal point not part of the corpus callosum. Voxel angle was set to be tangential to the corpus callosum. A semi-LASER^1^H-MRS sequence (TR = 7500 ms, TE = 100 ms, bandwidth = 6000 Hz, N = 2048) was used to acquire 32 channel-combined, VAPOR [42] water-suppressed spectra as well as a water-unsuppressed spectrum to be used for spectral editing and quantification. All participants were asked to fix their gaze on a white cross (50% gray background) during MRS acquisition.

**Figure 2.**
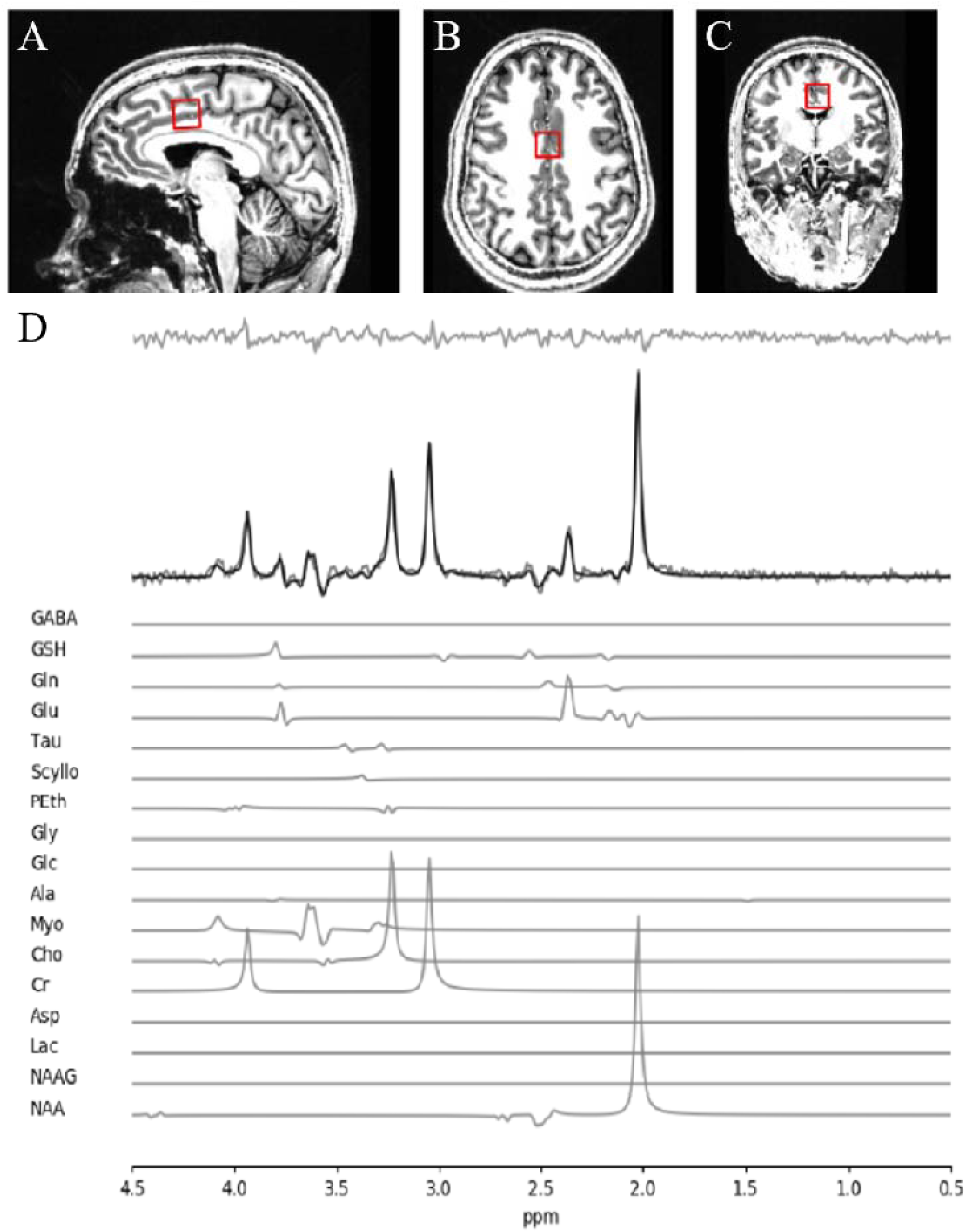
MRS voxel and spectra. (A) Sagittal, (B) axial, and (C) coronal view of voxel positioning on the dorsal anterior cingulate cortex. (D) Sample spectra obtained from a single volunteer. The bolded black line represents the fitted spectra with the residuals above and each individual metabolite contributions below.

Using the techniques outlined in Near and colleagues [43], the 32 spectra were phase and frequency corrected before being averaged into a single spectrum to be used for all subsequent analyses. QUECC [44] and HSVD [45] were applied to the spectrum for lineshape deconvolution and removal of residual water signal, respectively. Spectral fitting was done using fitMAN [46], a time-domain fitting algorithm that uses a non-linear, iterative Levenberg-Marquardt minimization algorithm to echo time-specific prior knowledge templates. The metabolite fitting template included 17 brain metabolites: alanine, aspartate, choline, creatine, γ-aminobutyric acid (GABA), glucose, glutamate, glutamine, glutathione, glycine, lactate, myo-inositol, N-acetyl aspartate, N-acetyl aspartyl glutamate, phosphorylethanolamine, scyllo-inositol, and taurine. Due to the long echo time used, no significant macromolecular contribution was expected. Metabolite quantification was then performed using Barstool [47] with corrections made for tissue-specific (gray matter, white matter, CSF) T_1_ and T_2_ relaxation through partial volume segmentation calculations of voxels mapped onto T_1_-weighted images acquired using a 0.75 mm isotropic MP2RAGE sequence (TR = 6000 ms, TI_1_ = 800 ms, TI_2_ = 2700 ms, flip-angle 1 (α_1_) = 4°, flip-angle 2 (α_2_) = 5°, FOV = 350 mm × 263 mm × 350 mm, T_acq_ = 9 min 38 s, iPAT_PE_ = 3 and 6/8 partial k-space). All spectral fit underwent visual quality inspection as well as Cramer-Rao lower bounds (CRLB) assessment for each metabolite.

Quality of metabolite quantification was measured using CRLB percentages for both groups using a CRLB threshold < 30% for glutathione to determine inclusion toward further analyses, in line with our prior study [18]. Notably, the mean CRLB for these metabolites were over two-folds lower than the individual threshold percentages. There was no significant difference in CRLB between the clinical high-risk group or healthy controls for both metabolites being reported in this study. We present the concentration and CRLB of other metabolites in our fitting template, along with the two presently mentioned, in the Supplementary Material. A sample of fitted spectrum for a single participant is presented in Figure 2.

### Clinical Assessments

Symptom severity was measured using the scale of prodromal symptoms (SOPS), on the same day of the scan. We also quantified the overall social and occupational functioning at the time of first presentation using SOFAS [48], administered on the same day of the scanning. To determine cannabis use in the past six months, the Cannabis Abuse Screening Test (CAST) was used [49]. The CAST is a six-item Likert-scale self-report questionnaire which asks the participant about cannabis use and how it effects their daily activities and relationships. Scores range from 6 to 30, with higher scores indicating more cannabis use. To determine alcohol use in the past 6 months, the Alcohol Use Disorders Identification Test-Concise (AUDIT-C) [50] was used. The AUDIT-C is a three-item Likert-scale self-report questionnaire which asks the participant about alcohol use frequency and quantity. Scores range from 0 to 12, with higher scores indicating more alcohol use. Alcohol users and nonusers were classified by AUDIT-C scores of four or more and less than four, respectively. Lastly, nicotine use in the past six months was determined by the single item Fagerström Test for Nicotine Dependence and smoking index [51]. The Fagerström test indicates time to the first cigarette after waking, and the smoking index is calculated by the number of years regularly smoking × the number of cigarettes per day, divided by 20 cigarettes per pack. A lower Fagerström test value indicates more nicotine dependence, and a higher smoking index indicates more nicotine use. The 10-item Drug Abuse Screening Test (DAST-10) [52] was also employed for substances other than cannabis, alcohol and nicotine, though our cohort did not endorse any such use.

### Bayesian Analysis

We evaluated the association between the [GSH] and SOFAS, SOPS, and CAST in the CHR group and the relationship between [GSH] and SOFAS in the HC group by using a Bayesian Spearman test [53]. This approach relies on data augmentation via Metropolis-within-Gibbs sampling algorithm. Briefly, we assumed the rank data as a reflection of a latent (truncated) normal distribution which allowed us to use a conventional likelihood function. That is to say, the latent continuous scores would manifest as “degraded” rank values. Following this assumption, the data augmentation algorithm would yield samples from a truncated posterior distribution. Here we tested the null hypothesis that ρ= 0 versus the alternative hypothesis that ρ∼Uniform [-1,1] (i.e., following a uniform prior distribution). We drew 11,000 samples using a Markov chain Monte Carlo (MCMC) method using the ‘spearmanCorrelation.R’ function in R as specified by van Doorn and colleagues [53]. We report the Bayes factor relative to the null model (BF_10_). BF_10_ > 1.0 suggests evidence in support of the (alternative) hypothesis and vice versa. We also report the mode and the proportion of the posterior distribution (i.e., posterior proportion, PP) of estimated ρ (Rho) values differing from zero along with the 95% highest density interval of the most credible values (HDI).

We estimated the posterior distribution of the (estimated) between-group differences in CAST, AUDICT-C, and SOFAS scores by means of the generalized linear model (GLM) within the context of hierarchical Bayesian parameter estimation as follows,

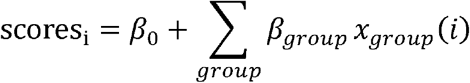

where the data conformed to a normal distribution around the predicted value (*score*) with a (wide) data-scaled uniform prior distribution for the standard deviation (s_i_). The baseline parameter (β_0_ had a data-scaled normal prior distribution with mean equal to the data mean and (wide) standard deviation relative to the standard deviation (SD_data_) of the data (1/(SD_data_ × 5)^2^). Group deflection parameters (*β*_*group*_ had normal prior distributions with mean zero and a Gamma prior distribution for the standard deviationσ_β_ with data-scaled shape and rate parameters (SD_data_/2 and 2 × SD_data_ respectively). This means thatσ_β_ provided informed priors on each group (deflection) parameter. In other words, groups would act as priors between each other. In total, we estimated posterior distributions of five free parameters (σ_i_, *β*_0_, *β*_*HC*_, *β*_*CHR*_, and σ_β_). Posteriors were estimated in RJAGS using MCMC, drawing 11,000 samples (thinning = 10). We report the PP of the between-groups difference in scores.

To evaluate for the effect of group after accounting for the effect of SOFAS scores, we included these scores as a covariate in the GLM,

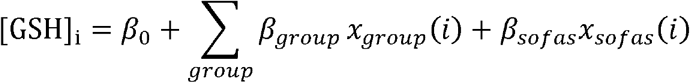

in which we added a normal prior distribution of the covariate parameter (*β*_*sofas*_) had zero mean and data-scaled standard deviation equal to 1/(2 × SD_GSG_data_ / SD_SOFAS_data_)^2^. In total, we estimated posterior distributions of six free parameters (σ_i_, *β*_*0*_, *β*_*HC*_, *β*_*HCR*_,*σ*_*β*_, and *β*_*sofas*_).

Posteriors were estimated in RJAGS using MCMC, drawing 11,000 samples (thinning = 10). We report the PP of the between-groups difference in [GSH] along with the 95% HDI. The posterior distribution of the effect size of this difference is also reported.

## Conclusion

In summary, our data offers preliminary support for GSH level as a clinically actionable marker of prognosis in emerging adults presenting with risk features for various severe mental illnesses. The use of a longitudinal approach to track GSH levels in future CHR studies may help establish the mechanistic primacy of antioxidant status in determining long term outcomes.

## Data Availability

All data is available upon request. Requests for data should be addressed to Dr. Lena Palaniyappan.

## Acknowledgements

We thank Mr. Trevor Szekeres, Mr. Scott Charlton, Mr. Joseph Gati for their assistance in data acquisition and archiving. We thank Dr. Rob Bartha and Dr. Dickson Wong for consultation provided on MRS analysis. We thank all research team members of the NIMI lab and all the staff members of the PEPP London team for their assistance in patient recruitment and supporting clinical care. We gratefully acknowledge the participants and their family members for their contributions. Requests for data should be addressed to Dr. Lena Palaniyappan.

## Supplementary Materials

### Metabolite Profiles

Out of 17 neurometabolites included in the spectral fitting template, eight metabolites passed the ≤ 45% individual CRLB threshold (Table S1).

**Table S1.**
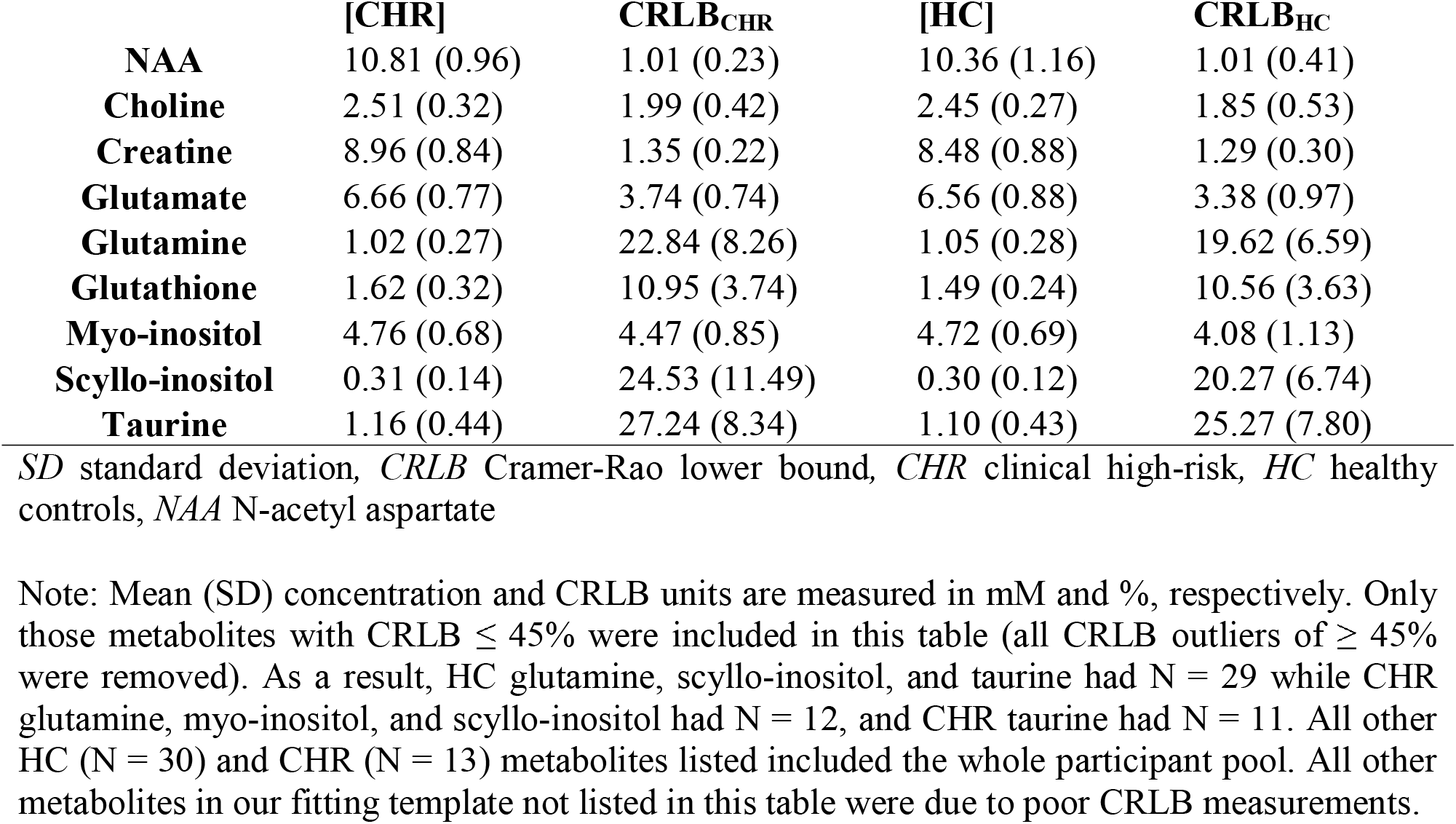
Mean metabolite concentration (SD) and mean CRLB (SD).

### Frequentist Analysis - Methods

All frequentist statistical tests were computed using IBM SPSS Statistics version 26 [1]. Group demographic differences were calculated using *t* tests and chi-square tests for continuous and dichotomous variables, respectively. Hierarchical regression was used to assess the effect of SOFAS and diagnosis (dummy coded CHR = 0, healthy controls = 1), with parameter estimates examined to test individual variable effects. Lastly, Spearman correlation was used to determine the correlation between metabolite levels and clinical scores (SOFAS, CAST, AUDIT-C, SOPS).

### Frequentist Analysis - Results

Upon further analysis using a frequentist approach with median splitting of glutathione concentrations in the patient group (Figure S1), a significant difference was found between SOFAS of low-glutathione (< 1.60mM) and high-glutathione (> 1.60mM) sub-groups (t(11) = - 2.49, *p* = 0.03), consistent with the Bayesian results relating GSH to SOFAS in CHR group.

**Figure S1.**
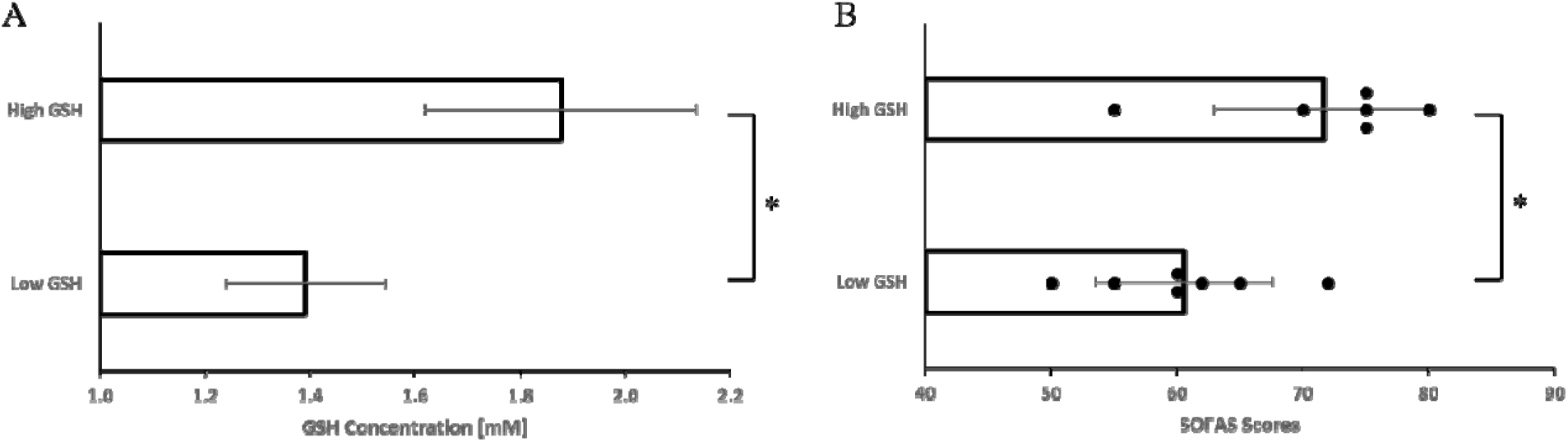
Median split analysis of glutathione (GSH) on SOFAS. (A) Mean (±SD) glutathione concentrations [mM] of low-GSH (< 1.60mM; N = 7) and high-GSH (> 1.60mM; N = 6) sub-groups. (B) Mean (±SD) SOFAS of the same low-GSH and high-GSH sub-groups with individual scores overlaid. Asterisk (*) denote significant difference between groups. In a hierarchical regression model with GSH level as the dependent variable, with SOFAS entered as a predictor for all subjects, the adjusted R^2^ of the model was -0.024, F(1, 41)□=□0.013, p□=□0.9, an insignificant effect. When CHR status was included in the model, the R^2^ of the model increased to 0.07. This R^2^ increase was statistically significant, F(1,40)□=□5.18, p□=□0.028. The regression coefficient for CHR status was significantly negative (B = -0.56, t = -2.28), indicating that GSH level was significantly higher in CHR subjects than in healthy controls after controlling for variance due to SOFAS. In this model, SOFAS had a trend level of significance as a predictor, indicating that higher SOFAS scores are seen in the presence of higher GSH levels (B = 0.43, t = 1.75, p = 0.09). These results are in keeping with the Bayesian analysis reported in the manuscript.

