## Supplementary material for "Glutathione as a molecular marker of functional impairment in patients with at-risk mental state: 7-Tesla ^1^H-MRS study": Table S1, Figure S1

Supplementary Materials

**Metabolite Profiles**

Out of 17 neurometabolites included in the spectral fitting template, eight metabolites passed the ≤ 45% individual CRLB threshold (Table S1).

| **Table S1.** Mean metabolite concentration (SD) and mean CRLB (SD). | | | | |
| --- | --- | --- | --- | --- |
|  | **[CHR]** | **CRLB_CHR_** | **[HC]** | **CRLB_HC_** |
| **NAA** | 10.81 (0.96) | 1.01 (0.23) | 10.36 (1.16) | 1.01 (0.41) |
| **Choline** | 2.51 (0.32) | 1.99 (0.42) | 2.45 (0.27) | 1.85 (0.53) |
| **Creatine** | 8.96 (0.84) | 1.35 (0.22) | 8.48 (0.88) | 1.29 (0.30) |
| **Glutamate** | 6.66 (0.77) | 3.74 (0.74) | 6.56 (0.88) | 3.38 (0.97) |
| **Glutamine** | 1.02 (0.27) | 22.84 (8.26) | 1.05 (0.28) | 19.62 (6.59) |
| **Glutathione** | 1.62 (0.32) | 10.95 (3.74) | 1.49 (0.24) | 10.56 (3.63) |
| **Myo-inositol** | 4.76 (0.68) | 4.47 (0.85) | 4.72 (0.69) | 4.08 (1.13) |
| **Scyllo-inositol** | 0.31 (0.14) | 24.53 (11.49) | 0.30 (0.12) | 20.27 (6.74) |
| **Taurine** | 1.16 (0.44) | 27.24 (8.34) | 1.10 (0.43) | 25.27 (7.80) |
| *SD* standard deviation*, CRLB* Cramer-Rao lower bound*, CHR* clinical high-risk*, HC* healthy controls, *NAA* N-acetyl aspartate | | | | |
| Note: Mean (SD) concentration and CRLB units are measured in mM and %, respectively. Only those metabolites with CRLB ≤ 45% were included in this table (all CRLB outliers of ≥ 45% were removed). As a result, HC glutamine, scyllo-inositol, and taurine had N = 29 while CHR glutamine, myo-inositol, and scyllo-inositol had N = 12, and CHR taurine had N = 11. All other HC (N = 30) and CHR (N = 13) metabolites listed included the whole participant pool. All other metabolites in our fitting template not listed in this table were due to poor CRLB measurements. | | | | |

**Frequentist Analysis - Methods**

All frequentist statistical tests were computed using IBM SPSS Statistics version 26 [1]. Group demographic differences were calculated using *t* tests and chi-square tests for continuous and dichotomous variables, respectively. Hierarchical regression was used to assess the effect of SOFAS and diagnosis (dummy coded CHR = 0, healthy controls = 1), with parameter estimates examined to test individual variable effects. Lastly, Spearman correlation was used to determine the correlation between metabolite levels and clinical scores (SOFAS, CAST, AUDIT-C, SOPS).

**Frequentist Analysis - Results**

Upon further analysis using a frequentist approach with median splitting of glutathione concentrations in the patient group (Figure S1), a significant difference was found between SOFAS of low-glutathione (< 1.60mM) and high-glutathione (> 1.60mM) sub-groups (t(11) = -2.49, *p* = 0.03), consistent with the Bayesian results relating GSH to SOFAS in CHR group .

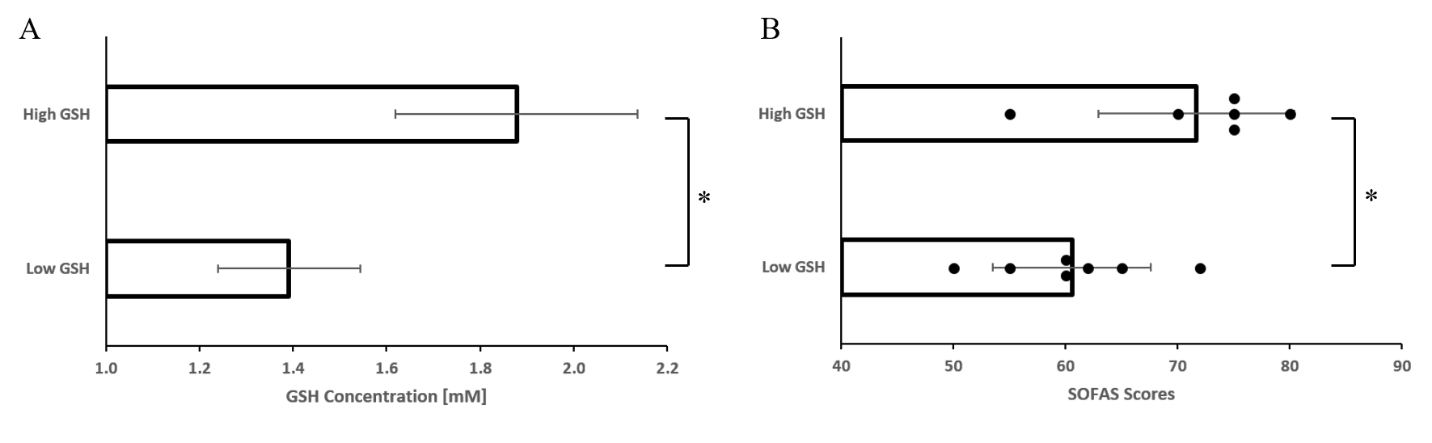

**Figure S1. Median split analysis of glutathione (GSH) on SOFAS.** (A) Mean (±SD) glutathione concentrations [mM] of low-GSH (< 1.60mM; N = 7) and high-GSH (> 1.60mM; N = 6) sub-groups. (B) Mean (±SD) SOFAS of the same low-GSH and high-GSH sub-groups with individual scores overlaid. Asterisk (*) denote significant difference between groups.

In a hierarchical regression model with GSH level as the dependent variable, with SOFAS entered as a predictor for all subjects, the adjusted R^2^ of the model was -0.024, F(1, 41) = 0.013, p = 0.9, an insignificant effect. When CHR status was included in the model, the R^2^ of the model increased to 0.07. This R^2^ increase was statistically significant, F(1,40) = 5.18, p = 0.028. The regression coefficient for CHR status was significantly negative (B = -0.56, t = -2.28), indicating that GSH level was significantly higher in CHR subjects than in healthy controls after controlling for variance due to SOFAS. In this model, SOFAS had a trend level of significance as a predictor, indicating that higher SOFAS scores are seen in the presence of higher GSH levels (B = 0.43, t = 1.75, p = 0.09). These results are in keeping with the Bayesian analysis reported in the manuscript.

References

1.	IBM Corp. IBM SPSS Statistics for Windows, Version 26.0. *2019* 2019.

Table Captions and Figure Legends

**Table S1.** Mean metabolite concentration (SD) and mean CRLB (SD).

**Figure S1. Median split analysis of glutathione (GSH) on SOFAS.** (A) Mean (±SD) glutathione concentrations [mM] of low-GSH (< 1.60mM; N = 7) and high-GSH (> 1.60mM; N = 6) sub-groups. (B) Mean (±SD) SOFAS of the same low-GSH and high-GSH sub-groups with individual scores overlaid. Asterisk (*) denote significant difference between groups.
